# Bacterial lipopolysaccharide as negative predictor of gemcitabine efficacy in advanced pancreatic cancer – translational results from the AIO-PK0104 phase 3 study

**DOI:** 10.1101/2020.05.01.20087668

**Authors:** Michael Guenther, Michael Haas, Volker Heinemann, Stephan Kruger, Christoph Benedikt Westphalen, Michael von Bergwelt-Baildon, Julia Mayerle, Jens Werner, Thomas Kirchner, Stefan Boeck, Steffen Ormanns

**Affiliations:** Institute of Pathology, Faculty of Medicine, Ludwig-Maximilians-University, Munich, Germany; Department of Internal Medicine III, Grosshadern University Hospital, Ludwig-Maximilians-University, Munich, Germany; German Cancer Consortium (DKTK), partner site Munich, Germany; Department of Internal Medicine II, Grosshadern University Hospital, Ludwig-Maximilians-University, Munich, Germany; Department of General, Visceral and Transplant Surgery, Ludwig-Maximilians-University, Munich, Germany

## Abstract

**Background:** Gram-negative bacteria mediated gemcitabine resistance in pre-clinical models. We determined if intratumoral lipopolysaccharide (LPS) detection by immunohistochemistry is associated with outcome in advanced pancreatic ductal adenocarcinoma (PDAC) treated with gemcitabine and non-gemcitabine containing 1^st^-line chemotherapy.

**Methods:** We examined LPS on tumor tissue from 130 patients treated within the randomized phase 3 trial AIO-PK0104 and a validation cohort (*n*=113) from a prospective biomarker study and analyzed the association of LPS detection to patient outcome according to treatment subgroups.

**Results:** In 24% of samples from the AIO-PK0104 study LPS was detected; in LPS-positive patients median OS was 4.4 months, compared to 7.3 months with LPS negative tumors (HR 1.732, *p*=0.010). A difference in OS was detected in the subgroup of patients treated with 1^st^-line gemcitabine-based treatment (n=71; HR 2.377, *p*=0.002), whereas no difference in OS was observed in the non-gemcitabine subgroup (n=59; HR 1.275, *p*=0.478). Within the validation cohort, the LPS positivity rate was 23%, and LPS detection was correlated with impaired OS in the gemcitabine subgroup (n=94; HR 1.993, *p*=0.008) whereas no difference in OS was observed in the non-gemcitabine subgroup (n=19; HR 2.596, *p*=0.219).

**Conclusions:** The detection of intratumoral LPS as a surrogate marker for gram-negative bacterial colonization may serve as a negative predictor for gemcitabine efficacy in advanced PDAC.

**Clinical trial registry:** NCT00440167

## Background

The majority of patients with pancreatic ductal adenocarcinoma (PDAC) is diagnosed in an advanced stage, resulting in mortality rates almost equal to incidence rates ^1^. Palliative chemotherapy is considered as an international standard of care for these patients; however, relevant predictive biomarkers for rational treatment decisions are still lacking ^2^. Increasing evidence indicates a crucial role for the human microbiome in solid malignancies, especially in gastrointestinal tumors which are in close contact to dense microbial colonization ^3, 4^. Importantly, intratumoral bacteria have been shown to promote tumorigenesis ^5^ and to confer resistance to gemcitabine in human adenocarcinoma ^6, 7^. As reported recently, intratumoral bacteria can metabolize gemcitabine (2′,2′-difluorodeoxycytidine) into its inactive form 2′,2′-difluorodeoxyuridine; in a pre-clinical model, gemcitabine depletion was dependent on the expression of a long isoform of the bacterial enzyme cytidine deaminase (CDD_L_) commonly found in gram-negative bacteria ^7^. Bacterial lipopolysaccharide is a major component of the cell wall of gram-negative bacteria and can easily be detected by immunohistochemistry^7^.

In the present study, we report the results of immunohistochemical detection of intratumoral LPS as a surrogate marker of gram-negative bacterial colonization in the tumor tissue of 243 patients with advanced PDAC from two independent study cohorts and the correlation of intratumoral LPS detection to patient outcome according to the applied 1^st^-line chemotherapy regimen (gemcitabine *vs* non-gemcitabine containing).

## Materials and Methods

### Patients and tumor samples

Archival tumor material was derived from 130 advanced PDAC patients treated within the randomized phase 3 study AIO-PK0104 (NCT00440167), that compared a 1^st^-line treatment with gemcitabine + erlotinib versus capecitabine + erlotinib (with the option of a cross-over to the comparator cytotoxic drug in the 2^nd^-line setting) ^8^. A validation of the LPS results from AIO-PK0104 was performed on 113 patient samples derived from a previously reported prospective biomarker trial ^9^, in which the 1^st^-line chemotherapy regimens were as follows: 40 patients received gemcitabine monotherapy, 29 patients gemcitabine and cisplatin, 10 patients gemcitabine and oxaliplatin, 14 gemcitabine and capecitabine, 11 patients single-agent capecitabine, 7 patients capecitabine and oxaliplatin, one patient received FOLFOX-6 and one patient received gemcitabine and 5-FU. Both studies had approval of the local ethics committee and were conducted according to GCP/ICH.

### LPS immunohistochemistry

Intratumoral bacterial LPS was detected immunohistochemically on formalin fixed paraffin embedded (FFPE) tumor material as described previously ^10^ using a monoclonal mouse anti-LPS antibody detecting the core bacterial LPS antigen (clone WN1 222-5, Hycult Biotech, Uden, The Netherlands) at a 1:800 dilution. Tumors from 133 patients were included in a tissue microarray (TMA) as three cores of one mm diameter each (71 samples from the AIO-PK0104 cohort, 62 samples from the validation cohort), whereas biopsies from 110 patients were stained as whole mount tissue sections (59 samples from the AIO-PK0104 cohort, 51 samples from the validation cohort). Normal human colonic mucosa, which physiologically carries high amounts of gram-negative bacteria and normal human liver tissue, which physiologically may show high amounts of LPS, were used as positive controls in each staining run (suppl. figure S1A and S1B). Of note, normal adjacent pancreatic tissue did not show LPS signals nor unspecific staining in acinar cells (figure S1C). On TMA embedded tumor tissue, cases showing strong signals in at least one TMA core or weak and single signals in at least two cores were evaluated as positive, whereas all others were evaluated as negative. On whole mount tissue sections, the presence of definitive LPS signals throughout the tumor tissue was evaluated as positive, whereas complete absence of signal was evaluated as negative. All slides were read and scored by two researchers (MG, SO), blinded to the patient outcome and discrepant cases were discussed until agreement was reached. Microphotographs were acquired as described previously ^10^.

### Statistical analyses

Progression free survival (PFS) and overall survival (OS) were calculated from the initiation of 1^st^-line chemotherapy to the occurrence of an event (progress, death). The correlation of tumor or patient characteristics to PFS and OS was calculated using the Kaplan-Meier method. Hazard ratios were estimated by Cox proportional hazards regression. Correlation of patient or sample characteristics was assessed using cross tabulations and two sided x2-tests. Statistical analyses were run on SPSS software (IBM, Ehningen, Germany).

## Results

### Patient characteristics

In the overall population (n=243) median patient age was 62 years; 145 patients were male. On therapy initiation, 23 patients from the AIO-PK0104 trial and 10 patients from the validation cohort had locally advanced disease, whereas all others had distant metastases. Both cohorts were comparable in terms of patient age, sex, KPS groups as well as tissue sample origin and tissue modality (TMA vs whole slide). In the AIO-PK0104 cohort, we detected more locally advanced cases, higher grade tumors and less lung metastases. Related to the trial design, more patients received non-gemcitabine based 1^st^-line chemotherapy in the AIO-PK0104 cohort (table S2). All clinicopathologic patient characteristics and associated survival times of each subgroup are summarized in table S3.

### LPS detection and clinico-pathological patient characteristics

Using immunohistochemistry, bacterial LPS could reliably be detected in 24.5 % of all PDAC samples (figure 1). To preclude potential sampling errors due to the use of TMA embedded tissue, we compared the staining result in whole mount tissue sections and the TMA cores of 10 exemplary positive and 15 exemplary negative cases from both cohorts and found complete consistency (table S4, figures S5A, S5B). Interestingly, we detected a significantly higher rate of LPS positivity in tumor samples which were derived from metastatic sites compared to primary tumor tissue samples (36.5% vs. 9.4%, p<0.001, table 1). As expected in a cohort of advanced PDAC patients, tissue samples from liver metastases constituted the largest group (96 of 243 samples) and showed a significantly higher LPS positivity rate than tissue from other metastatic sites or primary tumors (55.2% vs. 9.3%, p<0.001, table 1). In the subgroup in which only primary tumor samples were examined, no statistically significant association of primary tumor location and LPS detection was found (p=0.484, table S6). Moreover, we found no statistically significant association between LPS detection and patient characteristics such as gender, age group, KPS group or tumor differentiation (table 1).

**Figure 1.**
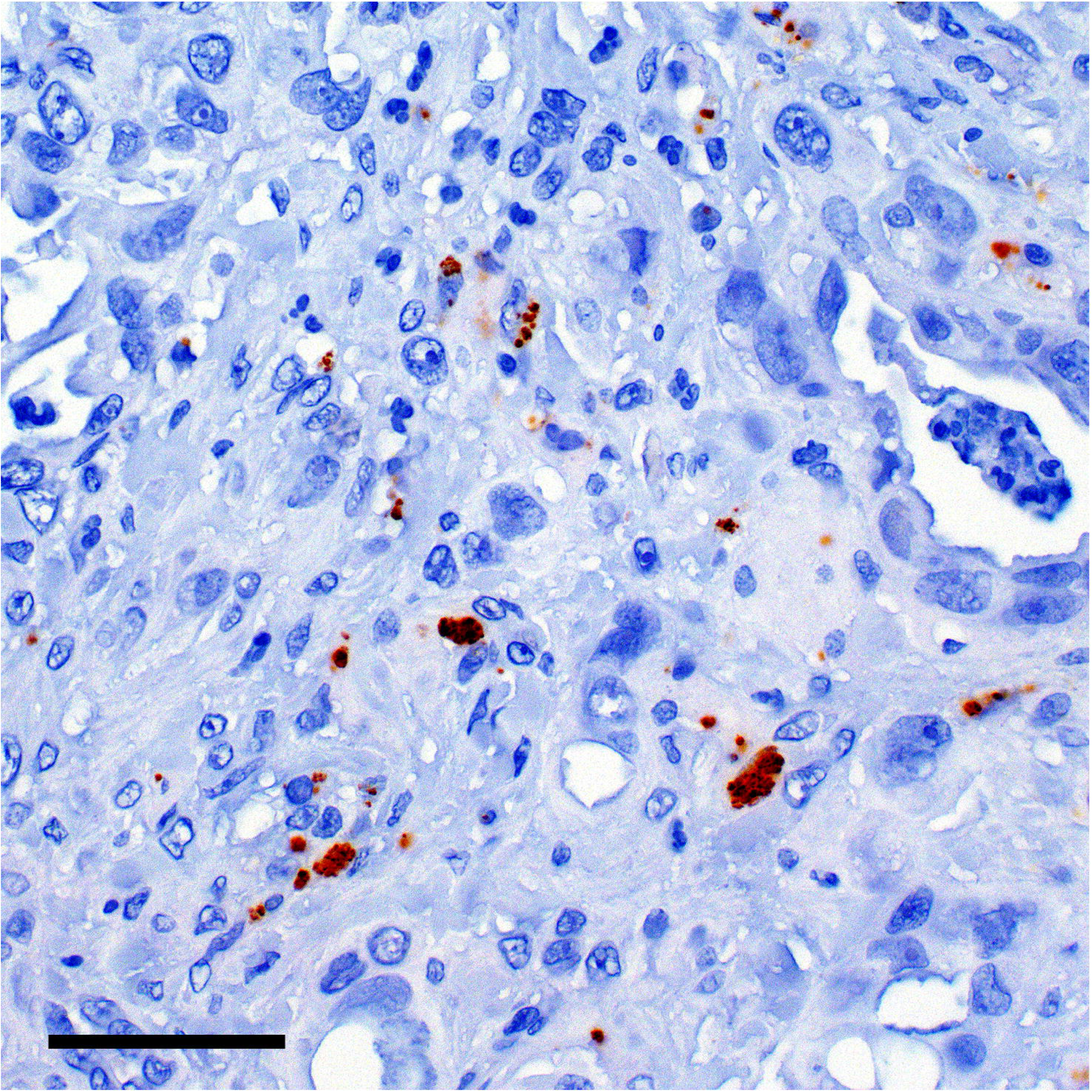
Intratumoral LPS can be detected in advanced PDAC tumor samples. Immunohistochemical detection of bacterial lipopolysaccharide (LPS) in pancreatic cancer. Exemplary cases of LPS positive (A) and LPS negative (B) pancreatic cancer samples. 200-fold magnification. Scale bars indicate 50 µm.

**Table 1.** Association of clinicopathologic variables and LPS detection in the entire study cohort.

| table 1 |  | LPS |  |  | p-value (χ<sup>2</sup>-test) |
| --- | --- | --- | --- | --- | --- |
|  |  | - | + | total |  |
| primary tumor location | unknown | 17 | 6 | 23 | 0.617 |
|  | pancreatic head | 103 | 36 | 139 |  |
|  | pancreatic body | 39 | 8 | 47 |  |
|  | pancreatic tail | 27 | 7 | 34 |  |
| total |  | 186 | 57 | 243 |  |
| metastasis at therapy initiation | lung | 5 | 1 | 6 | 0.023 |
|  | liver | 65 | 31 | 96 |  |
|  | peritonuem | 12 | 0 | 12 |  |
|  | other | 16 | 7 | 23 |  |
|  | none | 32 | 1 | 33 |  |
|  | liver and peritoneum | 17 | 1 | 18 |  |
|  | lung and liver | 6 | 5 | 11 |  |
|  | liver and bone | 1 | 0 | 1 |  |
|  | lung and abdominal lymph nodes | 3 | 0 | 3 |  |
|  | liver and lymph nodes | 16 | 6 | 22 |  |
|  | liver, lung and lymph nodes | 3 | 1 | 4 |  |
|  | lymph nodes | 8 | 3 | 11 |  |
|  | liver and adrenal gland | 1 | 1 | 2 |  |
|  | lung, liver and bone | 1 | 0 | 1 |  |
| total |  | 186 | 57 | 243 |  |
| tissue origin | hepatic metastasis | 59 | 44 | 103 | 0.000 |
|  | primary tumor | 106 | 11 | 117 |  |
|  | peritoneal metastasis | 8 | 0 | 8 |  |
|  | lung metastasis | 5 | 1 | 6 |  |
|  | lymph node | 4 | 0 | 4 |  |
|  | other / unknown | 4 | 1 | 5 |  |
| total |  | 186 | 57 | 243 |  |
| age group | <=60 | 75 | 27 | 102 | 0.346 |
|  | >60 | 111 | 30 | 141 |  |
| total |  | 186 | 57 | 243 |  |
| gender | male | 112 | 33 | 145 | 0.755 |
|  | female | 74 | 24 | 98 |  |
| total |  | 186 | 57 | 243 |  |
| KPS group | <= 80 | 75 | 28 | 103 | 0.210 |
|  | > 80 | 110 | 28 | 138 |  |
| total |  | 185 | 56 | 241 |  |
| CTX type | gemcitabine 1<sup>st</sup> line subgroup | 64 | 14 | 78 | 0.164 |
|  | non-gemcitabine 1<sup>st</sup> line subgroup | 122 | 43 | 165 |  |
| total |  | 186 | 57 | 243 |  |
| disease stage at therapy initiati | locally advanced | 32 | 1 | 33 | 0.003 |
|  | metastatic | 154 | 56 | 210 |  |
| total |  | 186 | 57 | 243 |  |
| grade group | G1-G2 | 75 | 14 | 89 | 0.031 |
|  | G3-G4 | 111 | 43 | 154 |  |
| total |  | 186 | 57 | 243 |  |
| tissue origin | metastasis | 80 | 46 | 126 | 0.000 |
|  | primary tumor | 106 | 11 | 117 |  |
| total |  | 186 | 57 | 243 |  |

### Intratumoral LPS detection is associated with inferior survival in gemcitabine treated PDAC patients

We detected bacterial LPS in the tumor material of 24 % of the 130 patients from the AIO-PK0104 study, which was significantly associated to inferior OS (4.4 *vs* 7.3 months, HR 1.732, *p*=0.010, figure 2A). Intratumoral LPS was significantly associated with a shorter OS in the 1^st^-line gemcitabine randomization arm (3.3 *vs* 7.7 months, HR 2.377, *p*=0.002, figure 2B), whereas no such difference in the 1^st^-line capecitabine arm was detected (5.7 *vs* 6.7 months, HR 1.275, *p*=0.478, figure 2C). All 113 patients selected for the validation cohort received erlotinib-free, either gemcitabine-based or non-gemcitabine based 1^st^-line chemotherapy regimens ^9^; 23% of these tumor samples were LPS positive, which again was significantly associated to shorter patient PFS (4.1 *vs* 7.8 months, HR 1.760, *p*=0.028) and OS (6.2 vs 10.8 months, HR 1.880, *p*=0.009, figure 2D). In subgroup analyses, the negative impact of LPS detection on either PFS (HR 2.051, *p*=0.010) or OS (HR 1.993, *p*=0.008, figure 2E) was restricted to the gemcitabine subgroup, whereas no difference in PFS (HR 1292, *p*=0.742) or OS (HR 2.596, *p*=0.219, figure 2F) was detected for the non-gemcitabine subgroup with regard to LPS status, respectively (table 2).

**Figure 2.**
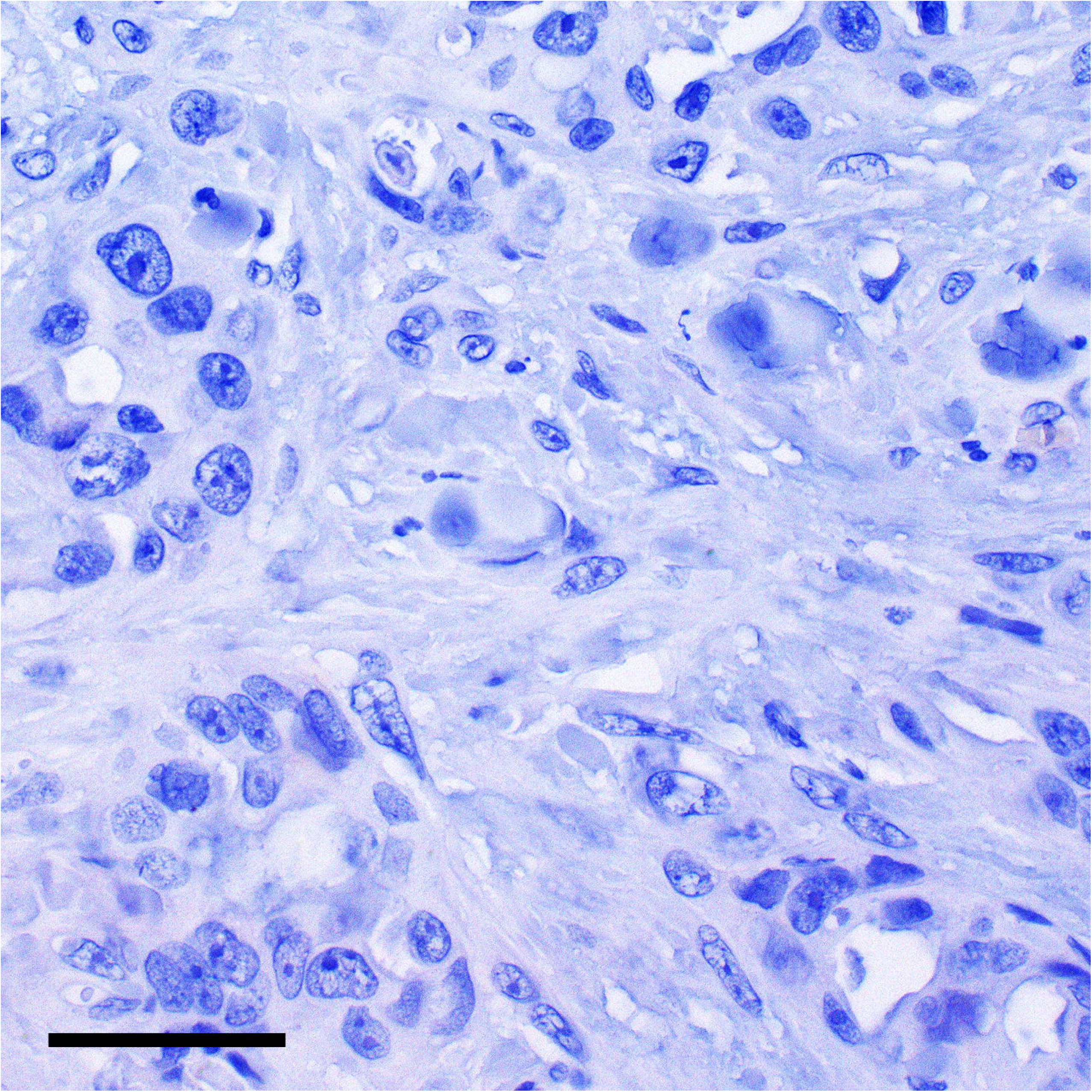
Intratumoral LPS is associated with inferior OS in gemcitabine treated PDAC patients. Overall survival (OS) of each patient subgroup according to intratumoral LPS detection. Kaplan-Meier plots of OS in the (A) AIO-PK0104 overall cohort (B) 1^st^-line gemcitabine subgroup of the AIO-PK0104 trial population (C) 1^st^-line capecitabine subgroup of the AIO-PK0104 trial population (D) Validation overall cohort (E) 1^st^-line gemcitabine subgroup of the validation cohort (F) 1^st^-line non-gemcitabine subgroup of the validation cohort Crossed lines indicate censored cases.

**Table 2.**
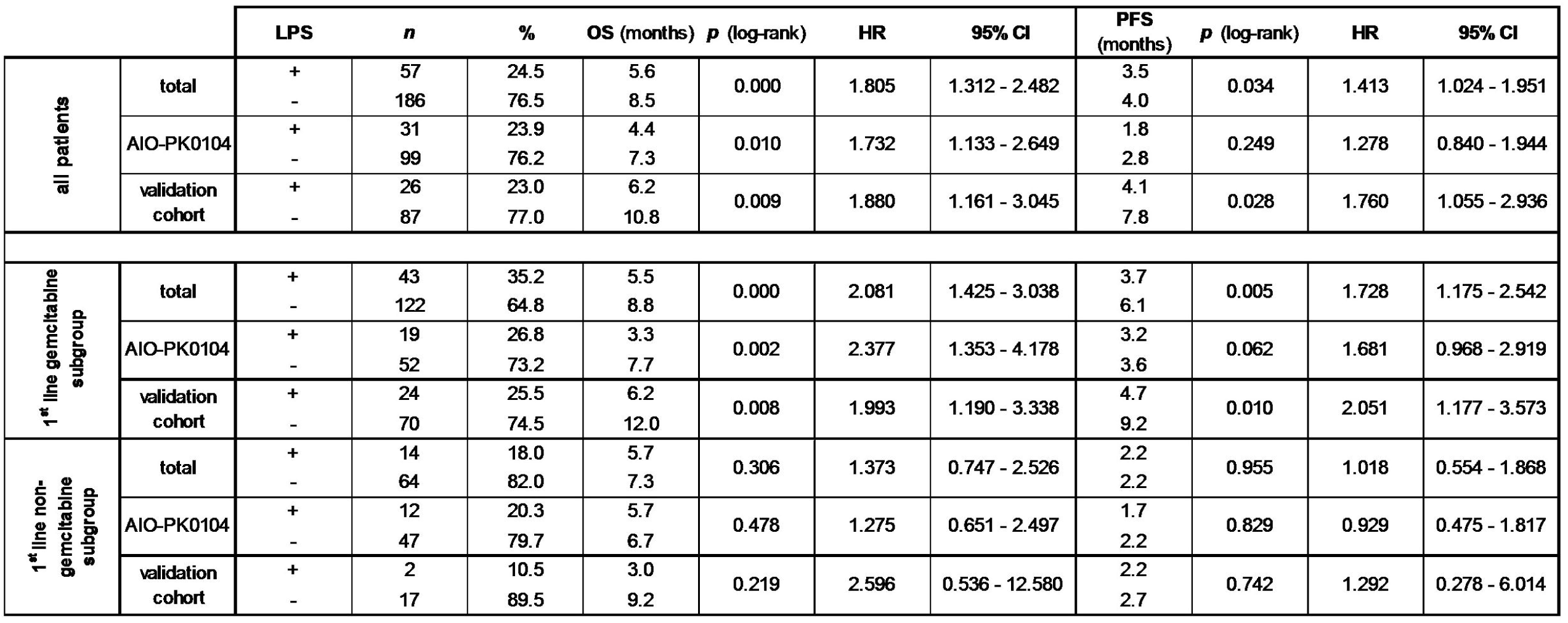
Patient prognosis according to LPS detection in each patient subgroup. Median times of overall survival (OS) and progression free survival (PFS) according to intratumoral LPS detection in each study subgroup as well as corresponding hazard ratios (HR) and 95% confidential intervals (95% CI).

In Cox multivariate regression analyses adjusting for the parameters which were statistically significant prognosticators in univariate analyses (tumor grade, KPS and disease stage at chemotherapy initiation), we confirmed LPS detection as independent negative predictor for both PFS and OS in the gemcitabine 1^st^-line chemotherapy subgroup (table 3).

**Table 3.** Intratumoral LPS is an independent negative prognostic biomarker in gemcitabine-treated PDAC patients. Cox regression analyses in the indicated patient subgroups for overall survival (OS) and (PFS), adjusting for tumor grade, KPS group, type of 1^st^-line palliative chemotherapy and disease stage at 1^st^-line therapy initiation. KPS: Karnofsky performance status; tumor grade: high grade (G3-G4) *vs* low grade (G1-G2) differentiation; CTX-type: type of 1^st^-line chemotherapy (gemcitabine-based *vs* non-gemcitabine based); disease stage: disease stage at 1^st^-line therapy initiation (metastatic vs locally advanced).

|  | **OS** |  |  |  |
| --- | --- | --- | --- | --- |
|  | **parameter** | **p (Cox)** | **HR** | **95% CI** |
| **all cases** | grade group | 0.002 | 1.592 | 1.194 - 2.122 |
|  | KPS group | 0.000 | 0.601 | 0.455 - 0.793 |
|  | LPS | 0.002 | 1.664 | 1.203 - 2.302 |
| **gemcitabine 1<sup>st</sup> line subgroup** | grade group | 0.000 | 1.977 | 1.401 - 2.790 |
|  | disease stage | 0.029 | 1.877 | 1.065 - 3-306 |
|  | LPS | 0.002 | 1.872 | 1.267 - 2.765 |
| **non-gemcitabine 1<sup>st</sup> line subgroup** | KPS group | 0.000 | 0.401 | 0.246 - 0.654 |

|  | **PFS** |  |  |  |
| --- | --- | --- | --- | --- |
|  | **parameter** | **p (Cox)** | **HR** | **95% CI** |
| **all cases** | grade group | 0.002 | 1.610 | 1.187 - 2.186 |
|  | KPS group | 0.002 | 0.637 | 0.476 - 0.853 |
|  | CTX type | 0.000 | 0.456 | 0.335 - 0.620 |
| **gemcitabine 1<sup>st</sup> line subgroup** | LPS | 0.009 | 1.674 | 1.136 - 2.467 |
|  | grade group | 0.001 | 1.838 | 1.275 - 2.648 |
| **non-gemcitabine 1<sup>st</sup> line subgroup** | KPS group | 0.002 | 0.435 | 0.258 - 0.733 |

To preclude a potential impact of the imbalance of LPS positivity in metastatic and primary tumor tissue, we compared OS and PFS according to LPS detection in each subgroup and obtained similar results as in the overall cohorts (table S7). From 110 patients treated within the AIO-PK0104 study, objective tumor response data (a secondary trial endpoint) was available. In the capecitabine 1 ^st-^line randomization arm, the disease control rate (DCR) of patients with LPS positive tumors was 40.0% (4 of 10 patients) compared to 42.5% (17 of 40 patients) in the patients with LPS negative tumors (p=0.589, table S8). In the gemcitabine 1^st^-line randomization arm, DCR of patients with LPS positive tumors was 66.7% (8 of 12 patients) compared to 62.5% (30 of 48 patients) for patients with LPS negative tumors (p=0.534, table S8). Thus, we detected no statistically significant differences in tumor response according to LPS detection and treatment arm.

## Discussion

In far contrast to other malignancies like colorectal or lung cancer, biomarkers that predict treatment efficacy of specific drugs are still lacking in PDAC. In 2017, Geller *et al* reported pre-clinical data suggesting that intratumoral bacteria mediate tumor resistance to gemcitabine ^7^. However, as to our best knowledge, these data have never been clinically validated yet. As gemcitabine is mainly applied in PDAC, we thought to validate the hypotheses from Geller and co-workers on patient samples from our previously conducted clinical trials in advanced PDAC ^8, 9^. As both the AIO-PK0104 cohort as well as the validation cohort contained patients treated with 1^st^-line gemcitabine and also non-gemcitabine containing chemotherapy, we thought to evaluate a potential predictive role for the occurrence of intratumoral bacteria on gemcitabine efficacy.

Unfortunately, no positivity rates of LPS immunohistochemistry were reported in the study by Geller et al., but the rate of 25% LPS positivity in our overall study population was significantly lower than the rate of 76% ‘bacterial DNA’-positive PDAC samples reported in their study ^7^. However, as their molecular assay detects all kinds of bacteria with superior sensitivity compared to immunohistochemistry and LPS is a cell wall component of gram-negative bacteria only, both figures are not directly comparable. One also might expect higher positivity rates examining whole mount primary tumor resection specimens - as performed by Geller and colleagues - compared to biopsy tissue or TMA embedded tumor material. Nevertheless, we precluded a potential bias resulting from tissue modality and confirmed the reliability of TMA embedded tissue to detect intratumoral LPS compared to whole mount tissue. Moreover, although our study population consisted of patients with advanced PDAC, where histological confirmation is often performed by a percutaneous biopsy of metastases ^9^, we even detected higher positivity rates in metastases than in primary tumors, which was mostly due to more LPS positive liver metastases. However, intratumoral LPS detection remained a potential negative predictor of gemcitabine efficacy irrespective of the tissue origin subgroup.

Our main results (summarized within table 2) provide evidence that LPS detection in FFPE tumor samples may serve as a negative predictor for gemcitabine efficacy in advanced PDAC, thereby confirming for the first time the previously published pre-clinical data ^7^. This potential predictive biomarker for gemcitabine efficacy may be of special interest, as additive antibiotic treatment in LPS positive cases could at least theoretically be used to eliminate bacterial colonization and thus overcome bacterially mediated chemotherapy resistance as already shown *in-vivo* ^7^. Although gemcitabine monotherapy has been largely replaced by more efficient chemotherapy regimens in the clinically fit patient ^11^, it is still widely used in the clinical practice, especially in patients with significant co-morbidities ^12^, where adjuvant antibiotic treatment in LPS-positive tumors could increase the unfortunately still suboptimal treatment efficacy.

Our study has several limitations that have to be taken into account when interpreting the results: the design was retrospective and a relevant heterogeneity on the applied treatment regimens (specifically in the validation cohort) may lead to a potential bias. Additionally, in both cohorts the use of 2^nd^-line therapy (usually with the alternative cytotoxic backbone, e.g. 5-fluorouracil-based after gemcitabine and vice versa) may have an impact on the results, specifically for OS. However, as the rate of 2^nd^-line therapy was below 50% in AIO-PK0104, a significant impact of 2^nd^- and further-line treatments on our results should not be expected. It must also be noted that all patients included in this LPS analysis were treated before the introduction of FOLFIRINOX ^13^ or gemcitabine/nab-paclitaxel ^14^. Additionally, the LPS immunohistochemistry used only detects a cell wall component of gram-negative bacteria and does not allow assessment of bacterial viability or further characterization of the bacterial infiltrate. As both AIO-PK0104 as well as the prospective biomarker study (used for the validation cohort) unfortunately had not collected information about the frequency of endoscopic retrograde cholangiopancreatography (ERCP) including placement of bile duct stents, we are not able to provide data on a potential correlation of the ERCP procedure (and/or the occurrence of cholangitis and subsequent use of antibiotic) in our patient population – aspects that of course would be of additional scientific interest in interpreting the reported data.

Nevertheless, based on the results from AIO-PK0104 and the validation cohort, intratumoral bacterial LPS detection in PDAC samples may serve as a novel negative predictive biomarker for gemcitabine efficacy, confirming previous pre-clinical observations. These findings must be validated prospectively and should be addressed in the light that this biomarker also may be modifiable by the use of antibiotics.

## Data Availability

all supplemental data can be obtained upon request

## Additional Information

### Ethics approval and consent to participate

In both study cohorts, the local ethics committee approved the use of patient material for translational research purposes (project numbers 554-11 and 401-15). All patients gave written informed consent to participate in their respective study upon study enrollment. This study was performed in accordance with the Declaration of Helsinki.

### Data availability

All supplemental data of this manuscript can be found on the journals website.

### Authors’ contributions

MG and SO read and scored immunohistochemical stainings, performed statistical analyses and drafted the figures. SO and SB drafted the manuscript. TK read slides and provided material. MH, SK, CBW, VH, MvBB, JM, JW and SB provided clinical patient data and patient material. All authors read and reviewed the manuscript.

## Acknowledgments

The authors would like to thank all patients and their families, nurses, study coordinators and AIO investigators for their active participation in the clinical trials that contributed data to this translational biomarker study.

We also thank A. Sendelhofert and A. Heier for excellent technical assistance.

## Supplementary figures and tables

**Figure S1**

*Bacterial lipopolysaccharide (LPS) detection in (A) normal colonic mucosa, (B) normal liver and (C) normal acinar pancreatic tissue*

Immunohistochemical detection of bacterial lipopolysaccharide (LPS) in normal colonic mucosa shows distinct and strong a cloud like luminal staining, whereas it appears dot-like and speckeld in normal liver. Normal pancreatic tissue shows no staining. 200-fold magnification. Scale bars indicate 50 µm.

**Table S2**

*Comparison of the clinicopathologic parameters in the AIO-PK0104 and the validation cohort*

**Table S3**

*Patient prognosis according to clinicopathologic variables in each patient subgroup*

Median times of overall survival (OS) and progression free survival (PFS) according to clinicopathologic variables in each study subgroup as well as corresponding hazard ratios (HR) and 95% confidential intervals (95% CI).

**Table S4 / Figure S5**

*Complete concordance of LPS detection in TMA cores and corresponding whole mount tissue sections*

Comparison of the staining results in TMA tissue and whole mount tissue sections of ten exemplary positive and 15 exemplary negative PDAC cases, of which five negative cases are depicted in figure S5A and five positive cases in figure S5B.

**Table S6**

*LPS detection is not associated to primary tumor location*

Cross tabulation of LPS positivity in PDAC primary tumor samples according to the primary tumor location.

**Table S7**

*Similar effect of LPS detection on PFS and OS in metastatic and primary tumor tissue origin subgroups*

Median times of overall survival (OS) and progression free survival (PFS) according to intratumoral LPS detection in each subgroup according to tumor tissue origin (primary tumor vs metastatic tissue) as well as corresponding hazard ratios (HR) and 95% confidential intervals (95% CI).

**Table S8**

*LPS detection is not associated with tumor response in the AIO-PK0104 cohort*

Disease control rates (DCR) according to LPS positivity in each randomization arm of 110 patients from the AIO-PK0104 trial. PD = progressive disease, SD = stable disease, PR = partial response, CR = complete remission.

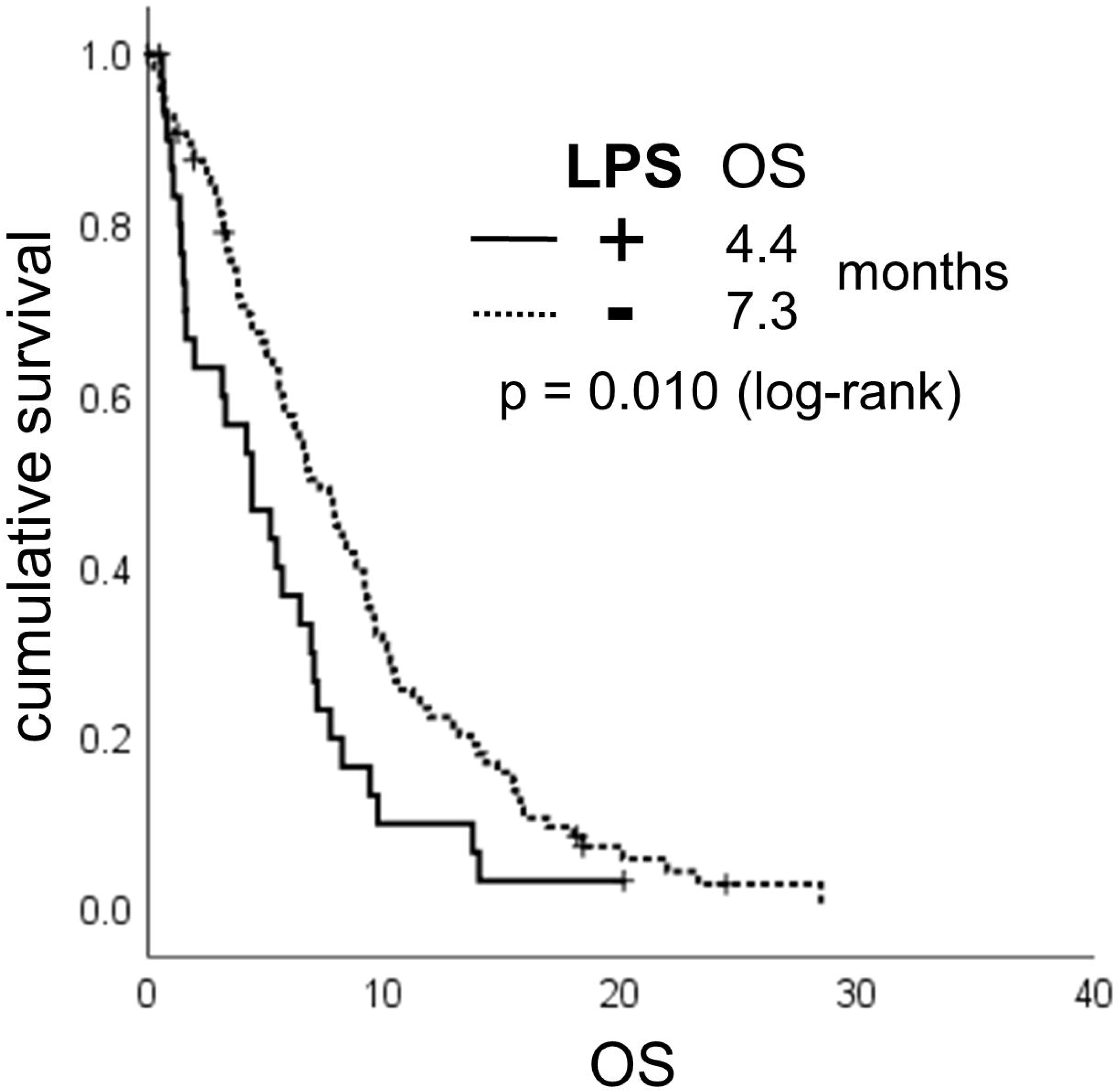

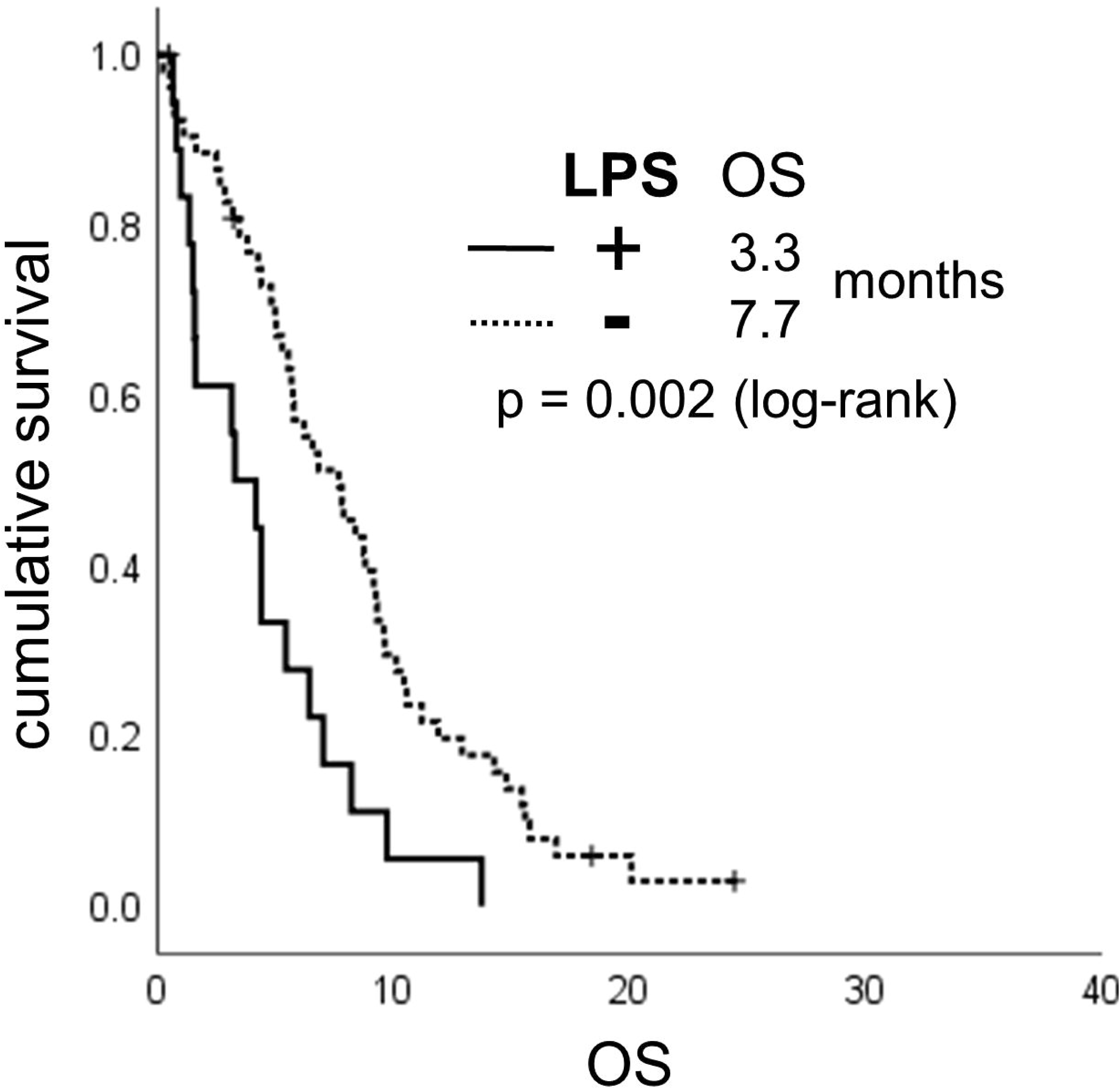

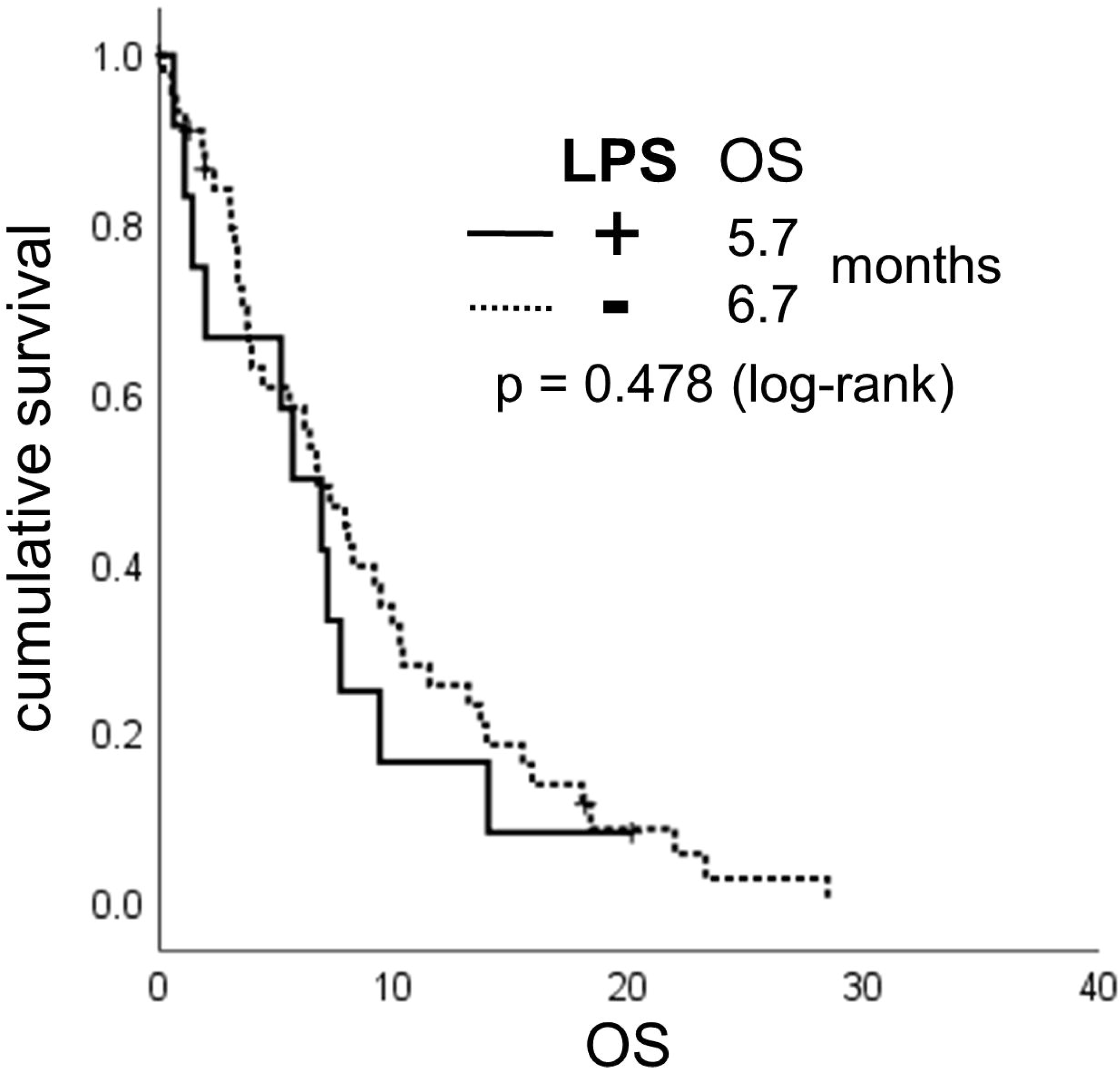

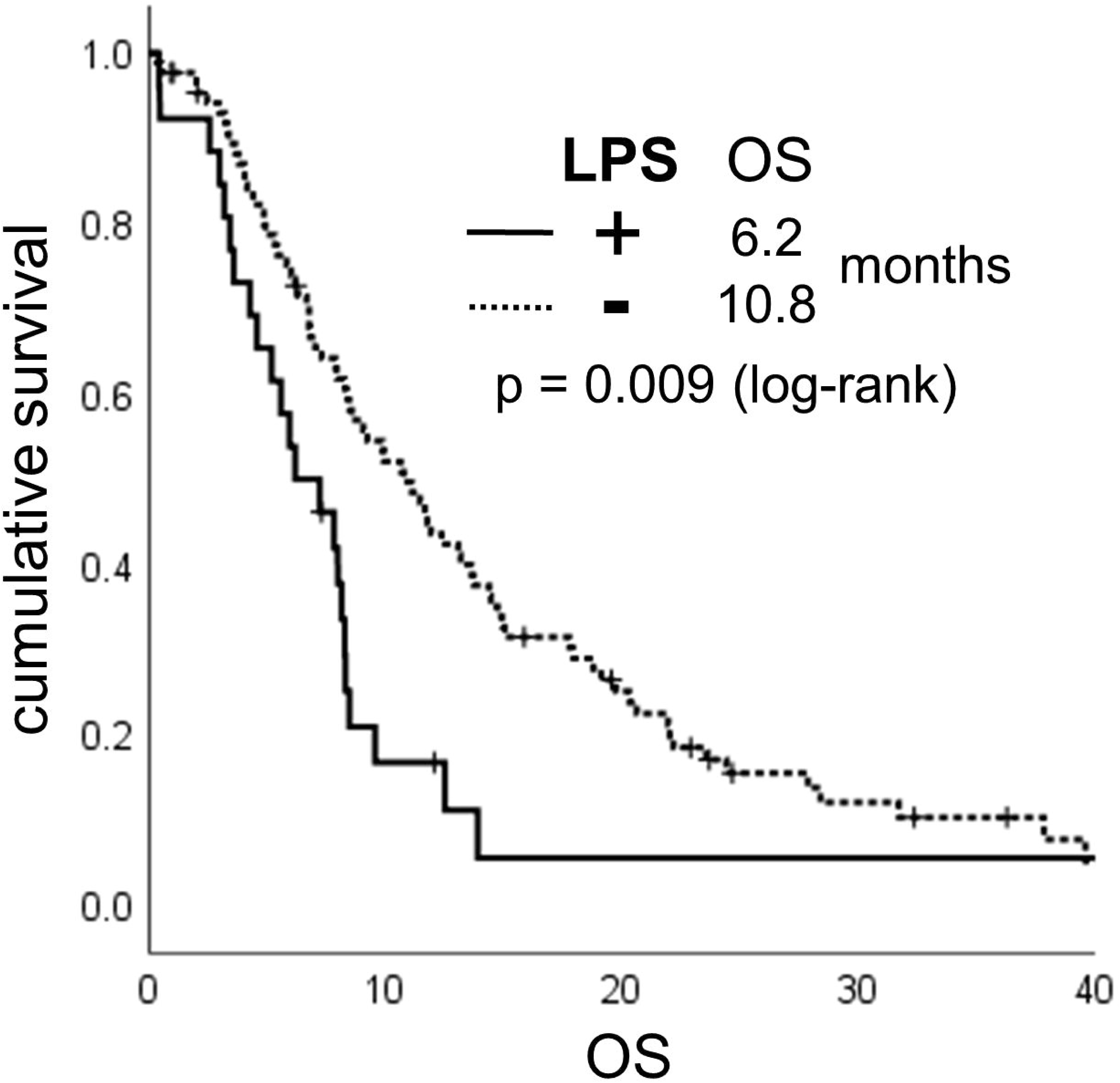

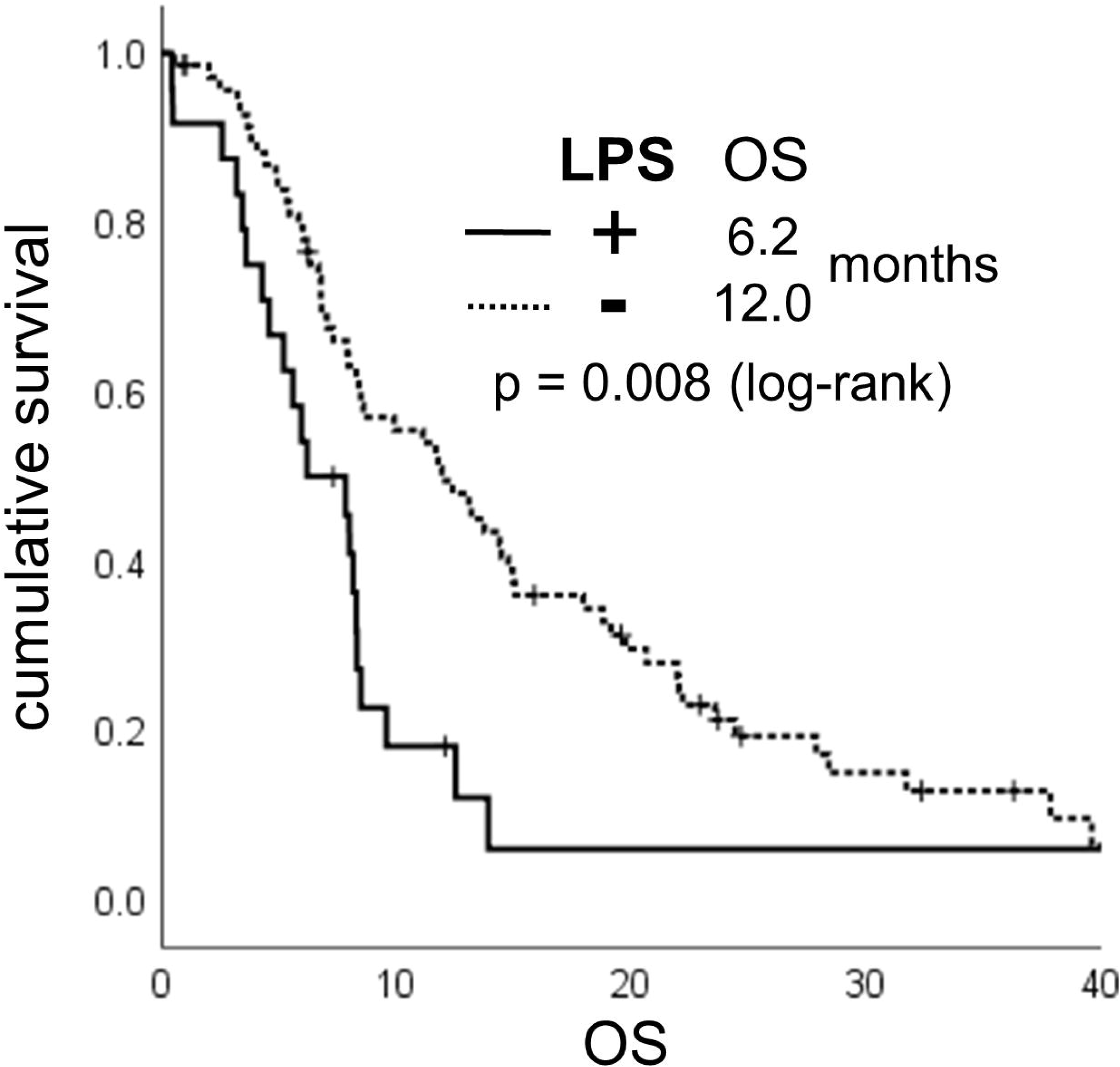

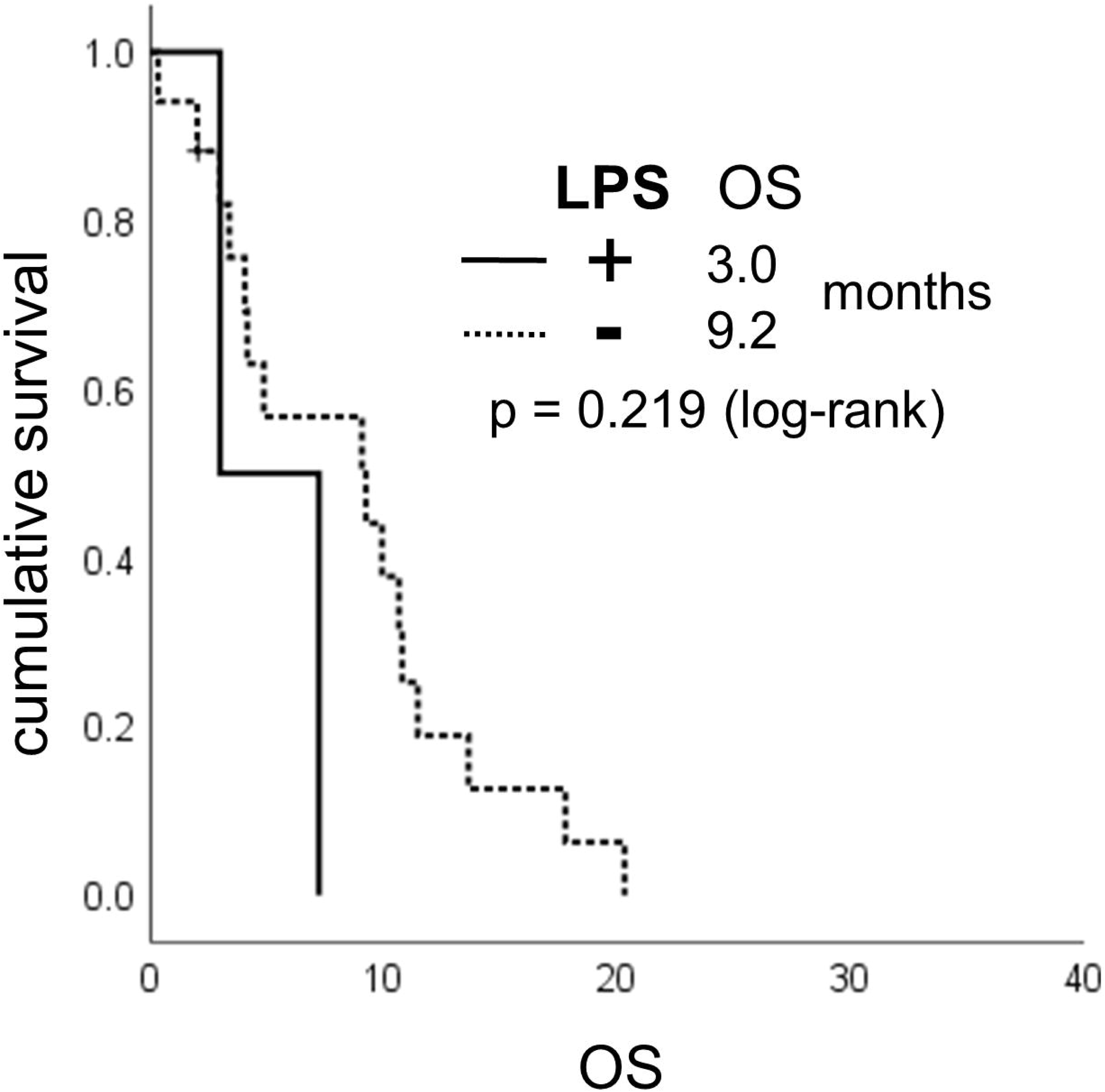

## References

1. Siegel RL, Miller KD, Jemal A. Cancer statistics, 2019. CA: a cancer journal for clinicians. 2019;69(1):7–34.

2. Neoptolemos JP, Kleeff J, Michl P, Costello E, Greenhalf W, Palmer DH. Therapeutic developments in pancreatic cancer: current and future perspectives. Nature reviews Gastroenterology & hepatology. 2018;15(6):333.

3. Riquelme E, Zhang Y, Zhang L, Montiel M, Zoltan M, Dong W, et al. Tumor microbiome diversity and composition influence pancreatic cancer outcomes. Cell. 2019;178(4):795–806. e12.

4. Helmink BA, Khan MW, Hermann A, Gopalakrishnan V, Wargo JA. The microbiome, cancer, and cancer therapy. Nature medicine. 2019: 1.

5. Sethi V, Kurtom S, Tarique M, Lavania S, Malchiodi Z, Hellmund L, et al. Gut microbiota promotes tumor growth in mice by modulating immune response. Gastroenterology. 2018;155(1):33–7. e6.

6. Lehouritis P, Cummins J, Stanton M, Murphy CT, McCarthy FO, Reid G, et al. Local bacteria affect the efficacy of chemotherapeutic drugs. Scientific reports. 2015;5:14554.

7. Geller LT, Barzily-Rokni M, Danino T, Jonas OH, Shental N, Nejman D, et al. Potential role of intratumor bacteria in mediating tumor resistance to the chemotherapeutic drug gemcitabine. Science. 2017;357(6356):1156–60.

8. Heinemann V, Vehling-Kaiser U, Waldschmidt D, Kettner E, Märten A, Winkelmann C, et al. Gemcitabine plus erlotinib followed by capecitabine versus capecitabine plus erlotinib followed by gemcitabine in advanced pancreatic cancer: final results of a randomised phase 3 trial of the ‘Arbeitsgemeinschaft Internistische Onkologie’ (AIO-PK0104). Gut. 2013;62(5):751–9.

9. Haas M, Ormanns S, Baechmann S, Remold A, Kruger S, Westphalen CB, et al. Extended RAS analysis and correlation with overall survival in advanced pancreatic cancer. British journal of cancer. 2017;116(11):1462.

10. Ormanns S, Altendorf-Hofmann A, Jackstadt R, Horst D, Assmann G, Zhao Y, et al. Desmogleins as prognostic biomarkers in resected pancreatic ductal adenocarcinoma. British journal of cancer. 2015;113(10):1460.

11. Springfeld C, Jäger D, Büchler MW, Strobel O, Hackert T, Palmer DH, et al. Chemotherapy for pancreatic cancer. La Presse Médicale. 2019;48(3):e159-e74.

12. Macarulla T, Carrato A, Díaz R, García A, Laquente B, Sastre J, et al. Management and supportive treatment of frail patients with metastatic pancreatic cancer. Journal of geriatric oncology. 2019;10(3):398–404.

13. Conroy T, Desseigne F, Ychou M, Bouché O, Guimbaud R, Bécouarn Y, et al. FOLFIRINOX versus gemcitabine for metastatic pancreatic cancer. New England Journal of Medicine. 2011;364(19):1817–25.

14. Von Hoff DD, Ervin T, Arena FP, Chiorean EG, Infante J, Moore M, et al. Increased survival in pancreatic cancer with nab-paclitaxel plus gemcitabine. New England Journal of Medicine. 2013;369(18):1691–703.

